# Discontinuity of the deadly infection rate for the COVID-19 pandemia due to lockdown measures

**DOI:** 10.1101/2020.08.05.20168880

**Authors:** Agustín Sabio Vera

## Abstract

An asymmetric version of the classical Kermack-McKendrick description of an epidemic evolution is presented in terms of four independent parameters. This is enough to obtain an accurate description of the different stages of the COVID-19 pandemia in any country for the reported daily and total number of casualties due to the infection. The asymmetry accounts for lockdown effects introduced to reduce the impact of the epidemic outburst. A set of new variables allows for an analytic study of the evolution of the system before and after the lockdown measures are put in place. A continuous matching is possible for all variables in the system apart from the time dependence of the infection rate. An analytic expression is obtained for this discontinuity which is proposed as a good quantity to gauge the efficiency of the lockdown measures. A study of this variable for different countries is performed.

## 1 An asymmetric Kermack-McKendrick model

For the mathematical treatment of a generic epidemic expansion a very well-known starting point is the classical work of Kermack and McKendrick in [1] where, among many other interesting results, they were able to describe the distribution of the number of deaths per day in a plague developed in Bombay from the end of 1905 until the summer of 1906. Their relevant formula and original graph are reproduced in Fig. 1.

**Figure 1:**
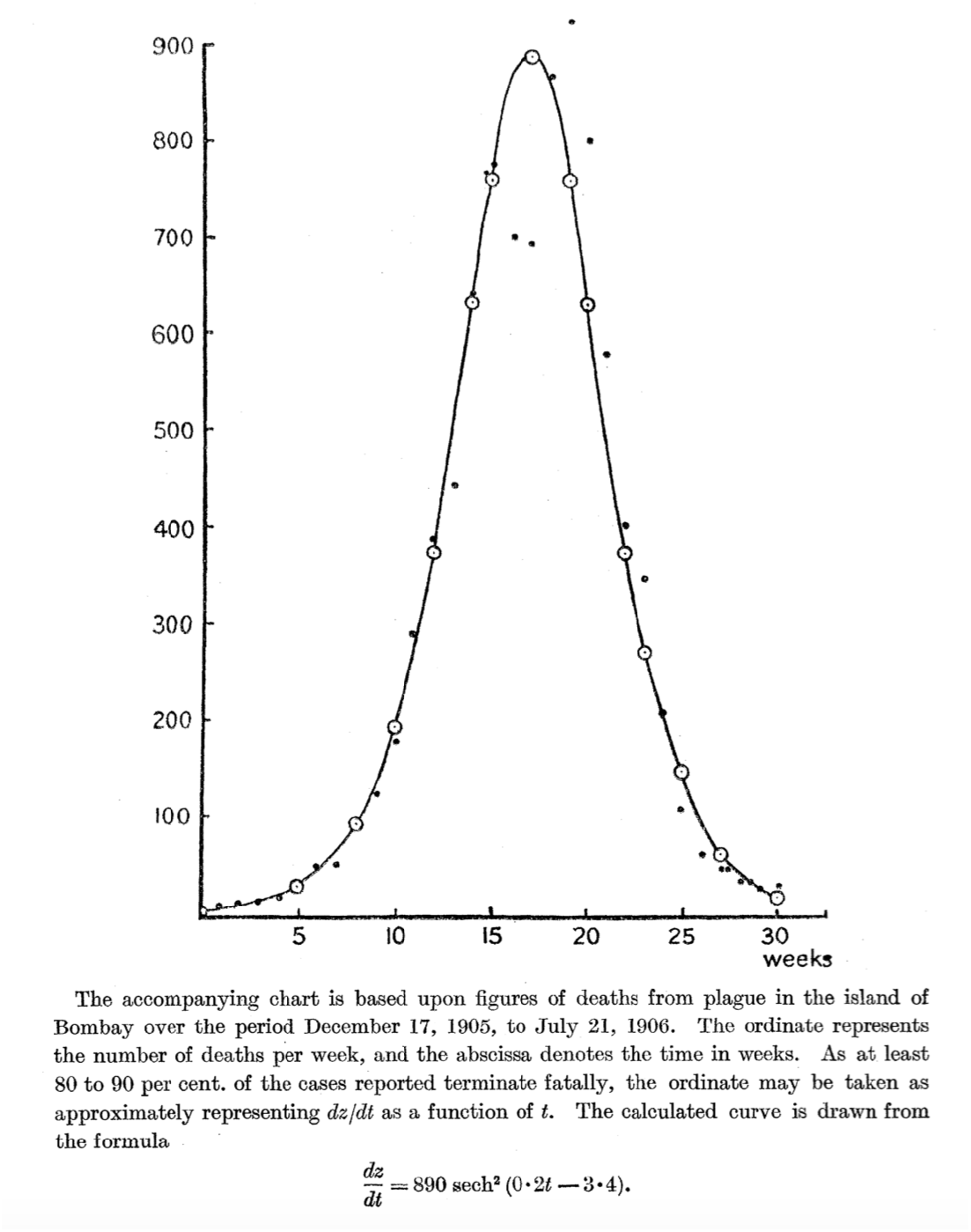
Original figure by Kermack and McKendrick in their 1927 paper 1.

Deterministic models for infectious disease epidemiology in their simplest formulation are based on three functions accounting for the number of individuals, *x*, susceptible to infection within a given population, the number of infected citizens, *y*, and those recovered, dead or isolated, *z*. Those in *x* can flow to *y* by infection at a rate proportional to *xy*. Elements from *y* can move to *z* by recovery, death or isolation at a rate proportional to *y*. The elements of *z* remain there and there is no possible entry into *x* (there exists a vast literature on these so-called SIR models, a review with some historical context can be found in, *e.g*., [2]). Kermack and McKendrick proposed the set of coupled differential equations [1]

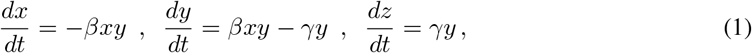

subject to the constraint *n* = *x* + *y* + *z*, where *n* corresponds to the subset of the total population exposed to the disease. To investigate the solution of this system of equations it is convenient to consider *γ* as constant with time and use a time rescalling of the form *τ* = *γt*, define 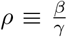 (also known as basic reproduction number) and write

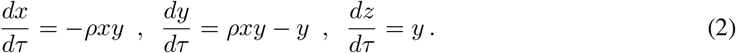

Following Kendall [3], the infection rate *ρ* = *ρ*(*z*) is not considered as a constant (there are many sophisticated models in this direction, see, *e.g*., [4–7]) since it can carry a time dependence through the function *z*(*τ*). Accounting for this dependence in *ρ* it is possible to write

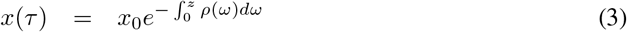

where *x*_0_ ≡ *x*(0). The classical approach is to consider *ρ*(*ω*) ≃ *ρ* = constant and approximate the solution to

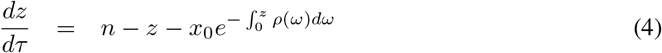

using a quadratic expansion of the exponential (an alternative to this expansion is to deal with delay equations [8]) to obtain

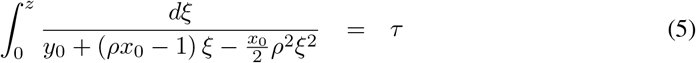

where *z*(0) = 0 and *y*_0_ ≡ *y*(0) = *n* − *x*_0_, as the fundamental equation to find *z*(*τ*). Kendall [3] pointed out that this widely used approximation in fact corresponds to the functional form

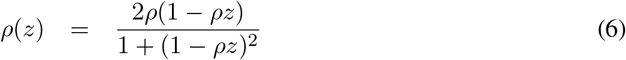

in Eq. (4). Taking this point of view in the present analysis, it is now useful to introduce the following change of notation for the parameters in the model:

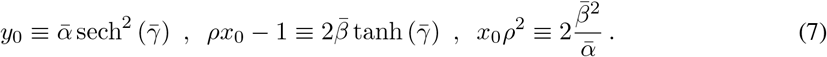

This allows to present Eq. (5) in the form

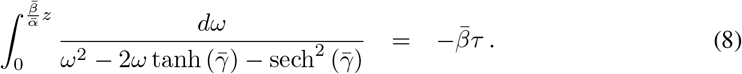

Its solution for *z* reads

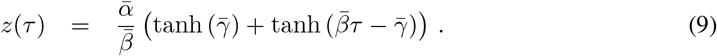

From Eq. (7) it is now possible to obtain

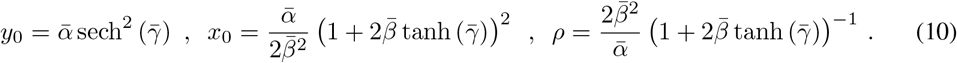

Using Eq. (6), the time dependence of the infection rate (note that there are many other approaches investigating this dependence with different methods, see, *e.g*., [9]) hence reads

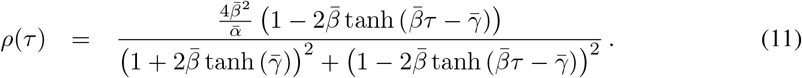

If, for simplicity, in *z*(*τ*) only the officially reported deaths due to the pandemia are included then the number of deaths per unit of time would be

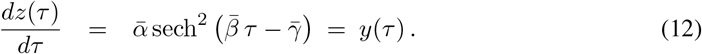

This differential distribution has a maximum value 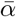 at the point 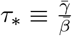 and it spans a total area

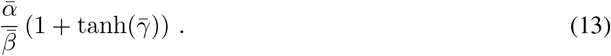

Relevant quantities at the point *τ*_*_ are

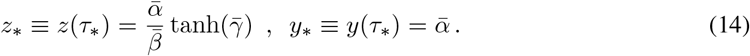

The solution in the region *τ* < *τ*_*_ for the complete system can therefore be written as

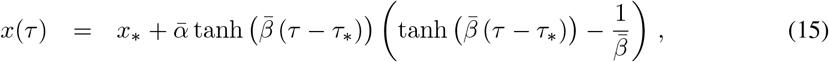

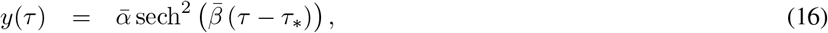

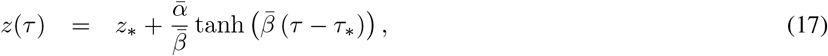

where

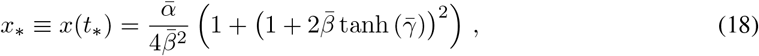

and

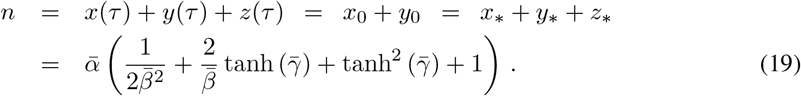

This procedure provides a description of the pandemia which is symmetric before and after *τ*_*_.

One of the main targets of the present work is to show that, in order to account for possible lockdown effects, after reaching the peak in *z*′ it is convenient to introduce an additional parameter, *δ*, in the form

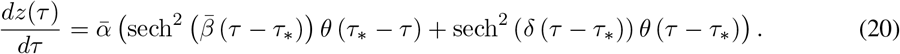

The corresponding integrated quantity up to time *τ* is

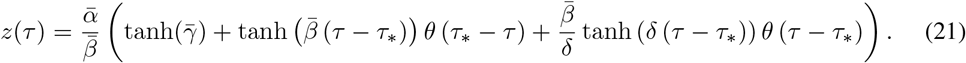

This generates an asymmetric description of the evolution of the system with a characteristic *shark-fin* shape in the *z*′ distribution. The symmetric flow is recovered when 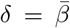. To fix *z*(*τ*) the relations

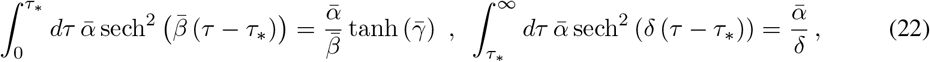

have been used. As a consequence

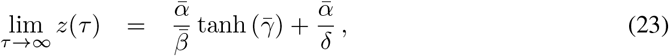

which is written as the sum of the number of total deaths before and after the peak of the *z*′ distribution.

In the region *τ* > *τ*_*_ there is a new system of equations where *ρ* is replaced by the function *η* (*γ* remains constant for all *τ*), *i.e*.

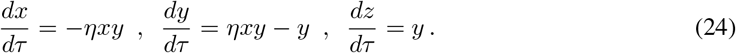

The initial conditions for the evolution must be set at *τ*_*_, hence

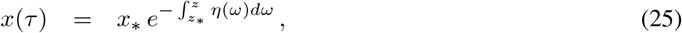

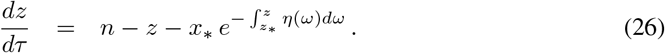

The *z* dependence for *η* in the form

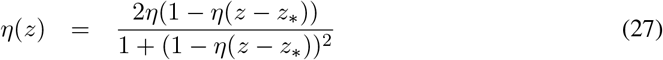

leads to

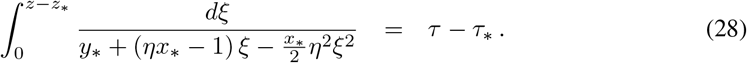

In this case the relations

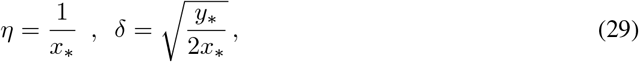

generate the solution for *τ* > *τ*_*_,

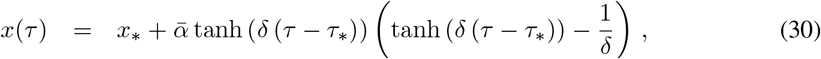

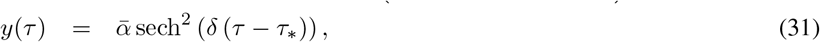

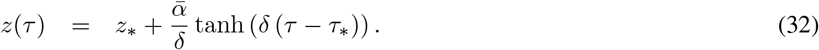

It can be checked that relation (19) also holds in this case. The expression for *δ* implies that

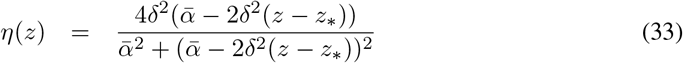

which in terms of time dependence reads

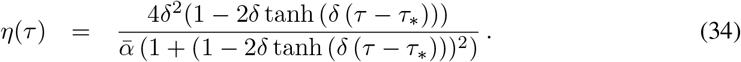

## 2 The COVID-19 pandemia in Spain

In order to test these expressions in a real scenario a study of the number of reported deaths due to COVID-19 fatal infections in Spain during 2020 is now discussed in some detail. In principle, the approach is simple. This subset of the total deaths during the pandemia is analysed, fitting the data to the previous description for *z*′(*τ*) and extracting the parameters 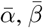 and 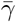 from it. It is important to note that this is a 3-parameter fit since *δ* is related to 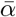, 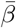 and 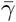 via Eq. (29)^1^.

The transition point *τ*_*_, which marks the peak of the *z*′ distribution, is also part of the fit since 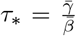. The first data point introduced in the analysis will correspond to that of the first day with a fatality associated to this pandemia in the country. To recall, the exact function used in the fit is Eq. (20) written as

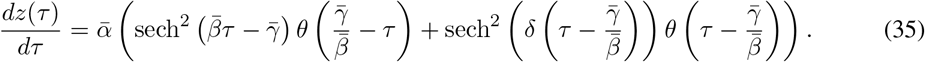

It turns out that the best fit under these conditions does not offer a precise description of *z*′. The obtained values for the independent parameters are

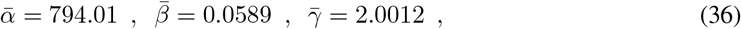

where, via Eq. (29), *δ* = 0.0557. This value is very close to 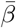 and the model still offers a description of the process too symmetric w.r.t. the maximum of *z*′. This can be seen in Fig. 2. The corresponding *x*, *y*, *z* solutions show an epidemic situation with a large susceptible population getting a small fraction of infected cases which finally die.

**Figure 2:**
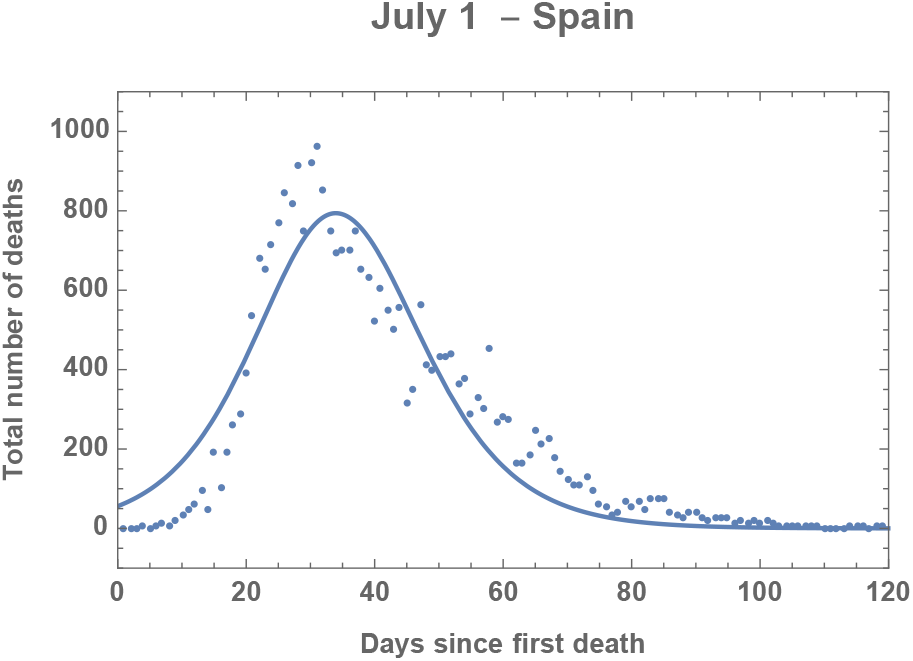
3-parameter fit to the data for daily number of deaths due to COVID-19 fatal infections in Spain.

An interesting feature of this type of 3-parameter fits is that the time dependence of the infection rate ρ for *τ* < *τ*^*^ and n for *τ* > *τ*^*^ is continuous. This can be seen in Fig. 3 where a very small initial infection rate evolves to a 0.9873 fraction of its value, small enough to stop the daily deaths. This continuity is mainly encoded in the relation (29).

**Figure 3:**
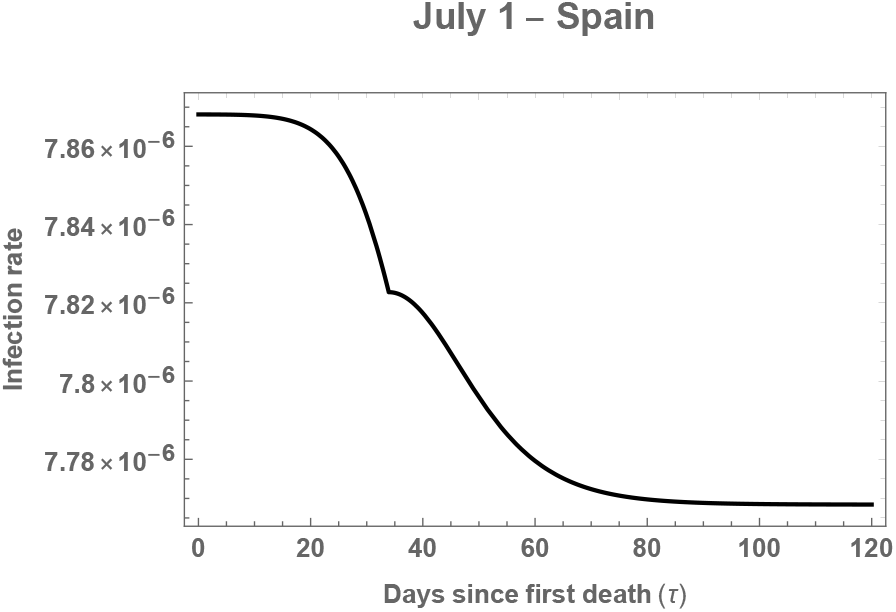
Time dependence of the infection rate given by the 3-parameter fit.

Clearly some of the constraints present in this analysis have to be relaxed in order to get a precise description of the process. The next step is then to try to fit the data with a 4-parameter function where *δ* is treated as an independent variable.

The comparison of the best 4-parameter fit to the data spanning 120 days of epidemic expansion in Spain (up to July 1) can be found in Fig. 4. The values of the different parameters are

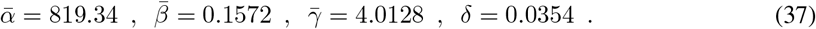

**Figure 4:**
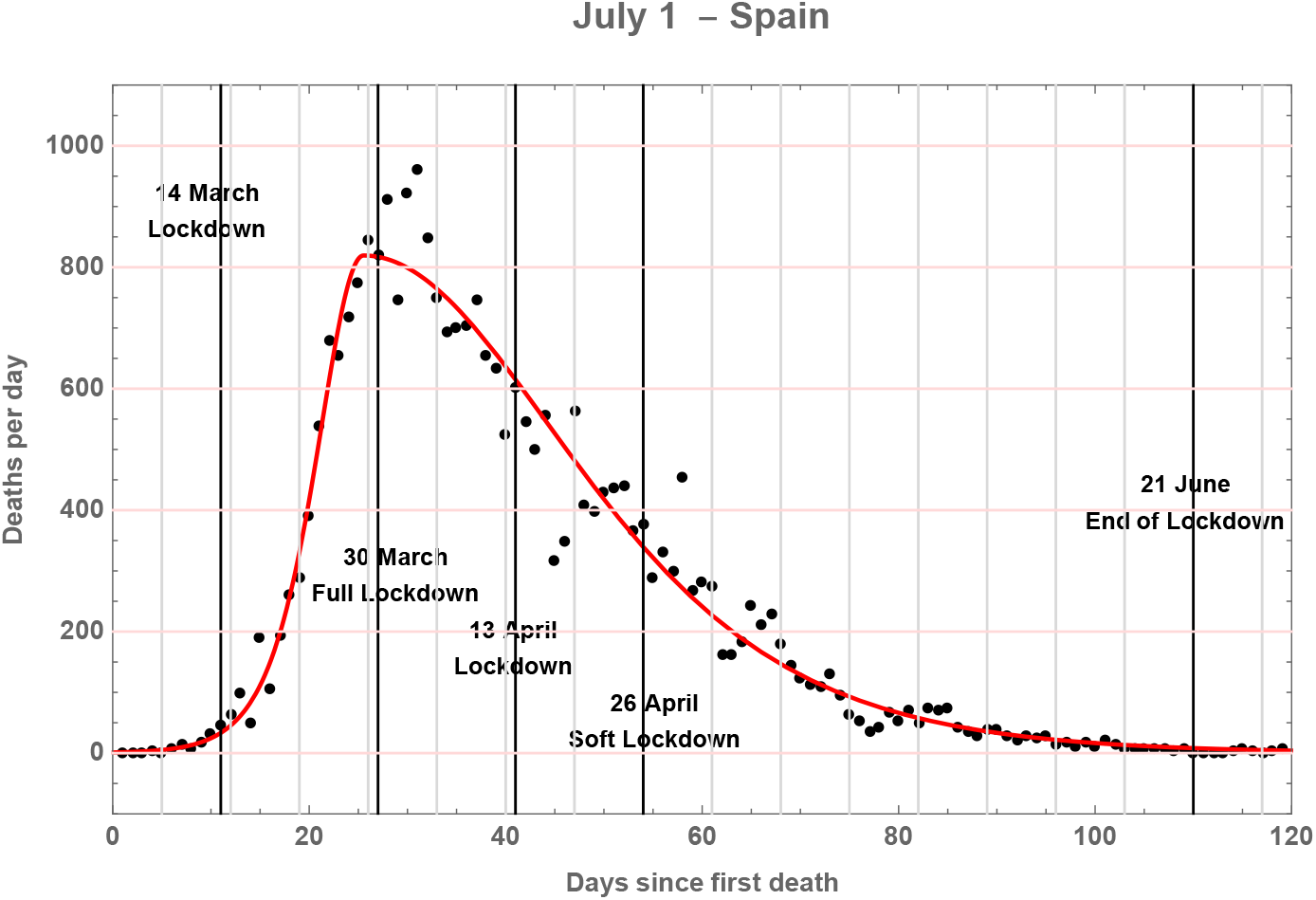
Fit to the data for the daily number of deaths due to COVID-19 fatal infections in Spain.

The figure manifests the characteristic shark-fin shape of the curve. The vertical lines indicate weekly periods and the dates when lockdown measures of different intensity were enforced. The same values for the parameters are introduced in Eq. (21) to describe the accumulated total number of deaths at each day in Fig. 5. It can be seen that the description of the pandemic evolution in terms of daily and total deaths is very accurate.

**Figure 5:**
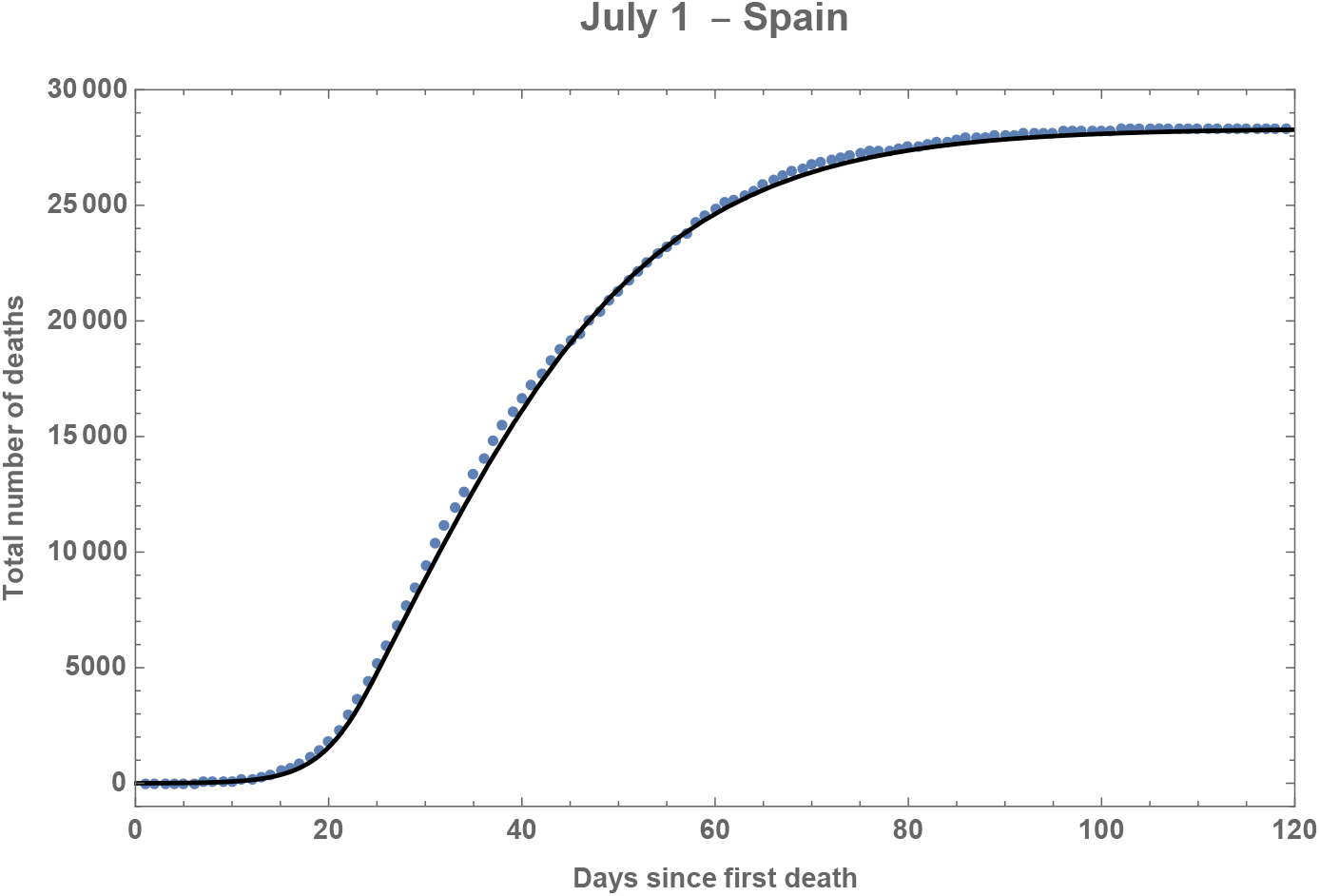
Fit to the data for the daily total sum of deaths due to COVID-19 fatal infections in Spain.

The functions *x*(*τ*), *y*(*τ*) and *z*(*τ*) are evaluated in Fig. 6. This is a more realistic representation of the situation under study which corresponds to a given number of the population where all susceptible individuals will suffer the disease and die. The continuity of the *x*, *y*, *z* functions is ensured by construction in the analytic expressions. This is not the case for the time dependence of the infection rate which is continuous only when Eq. (29) holds. When S is considered as the fourth free parameter, not necessarilly fulfilling Eq. (29), a discontinuity arises which will be a tangible effect of the lockdown measures. This point is studied in detail in the following.

**Figure 6:**
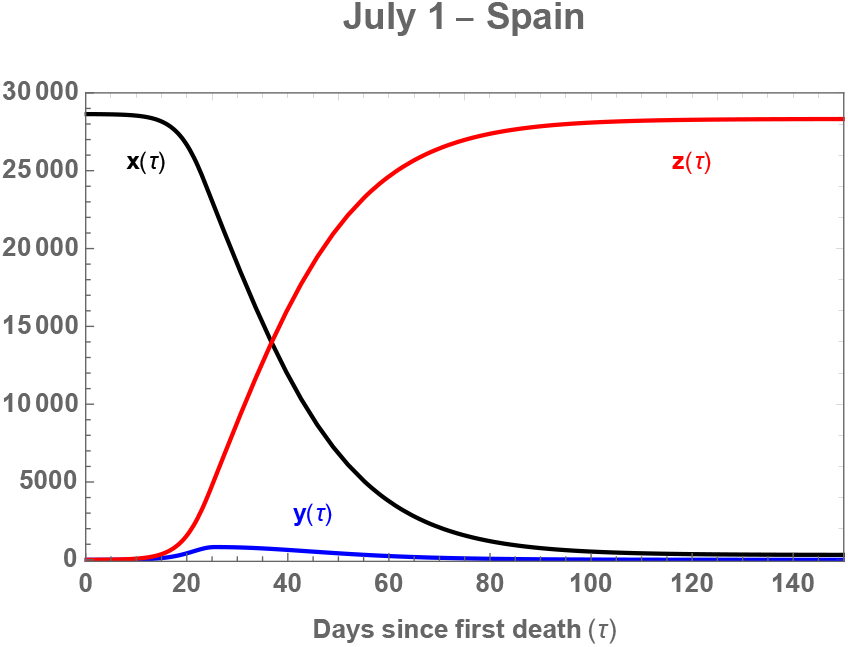
*x*, *y*, *z* solutions to the 4-parameter shark-fin model fit.

Eq. (29) would imply a value *δ* = 0.1346, much higher than the one obtained in the actual fit. The time dependence of the infection rate is investigated in two plots, Fig. 7 and Fig. 8, before and after the peak in the *z*′ distribution, respectively. They describe two quite distinct situations. For *τ* < *τ*^*^ the infection rate has a rather constant and high value of 4.58 · 10^−5^ which decreases fast when *τ* is close to *τ*^*^. The value at that point is *η*^*^ = *η*(*τ*^*^) =4.42 · 10^−5^. In the *τ* > *τ*^*^ region the value after 120 days is an order of magnitude smaller: 3.06 · 10^−6^. At that point the probability of infection is negligible, also at *τ* = *τ*^*^ where *ρ*^*^ = *ρ*(*τ*^*^) = 3.07 · 10^−6^.

**Figure 7:**
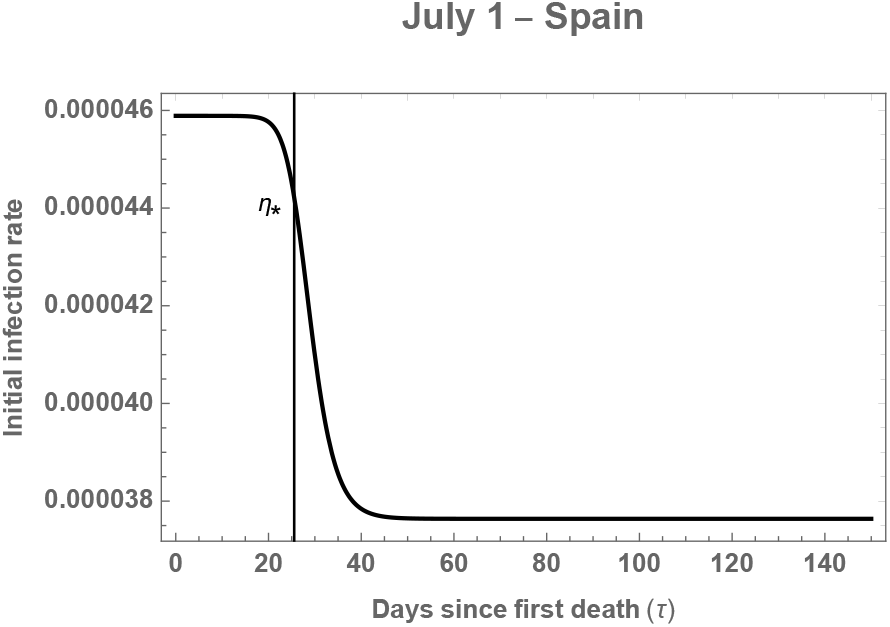
Time dependence of the infection rate given by the 4-parameter fit for *τ* < *τ*^*^.

**Figure 8:**
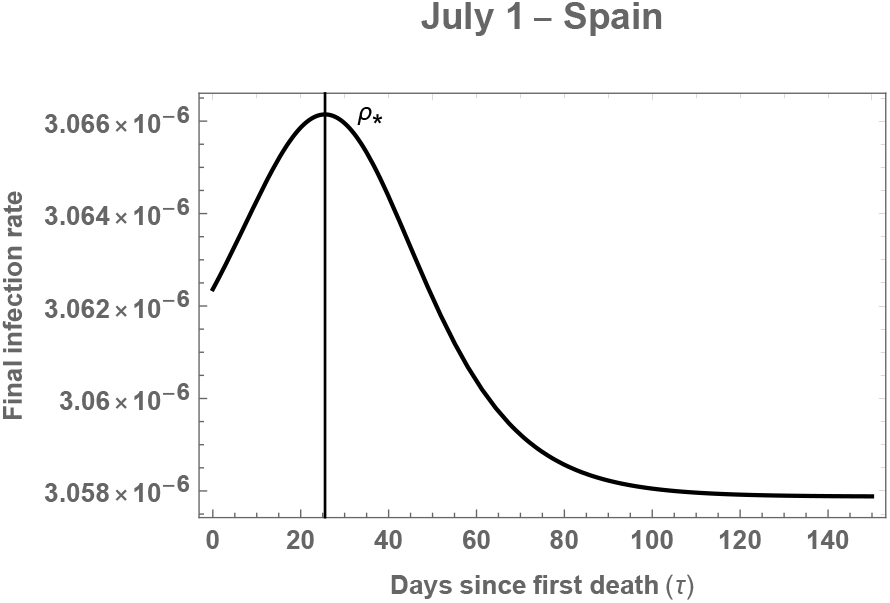
Time dependence of the infection rate given by the 4-parameter fit for *τ* < *τ*^*^.

In order to quantify this discontinuity it is useful to use the a-independent ratio

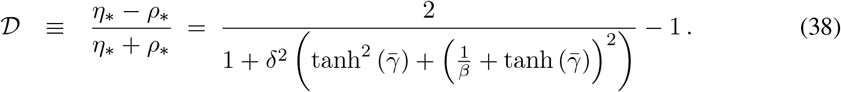

In the specific study of this section (after 120 days of pandemia) it is 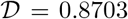. This quantity provides a measure of how effective a particular national lockdown was in order to reduce the infection rate. The closer it gets to one, the more efficient the lockdown measures are. Its value will be calculated for different countries in the next section.

Before doing so, a brief discussion of the predictive power of this approach is needed. The daily fit to the available data is shown in Fig. 9. The 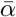, 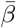, 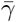 parameters reach quite accurate values already right after crossing the peak of *z*′. The *δ* parameter needs longer to reach convergence. The *τ*_*_ line shown in the curves corresponds to its value after 120 days of evolution in the system under study.

**Figure 9:**
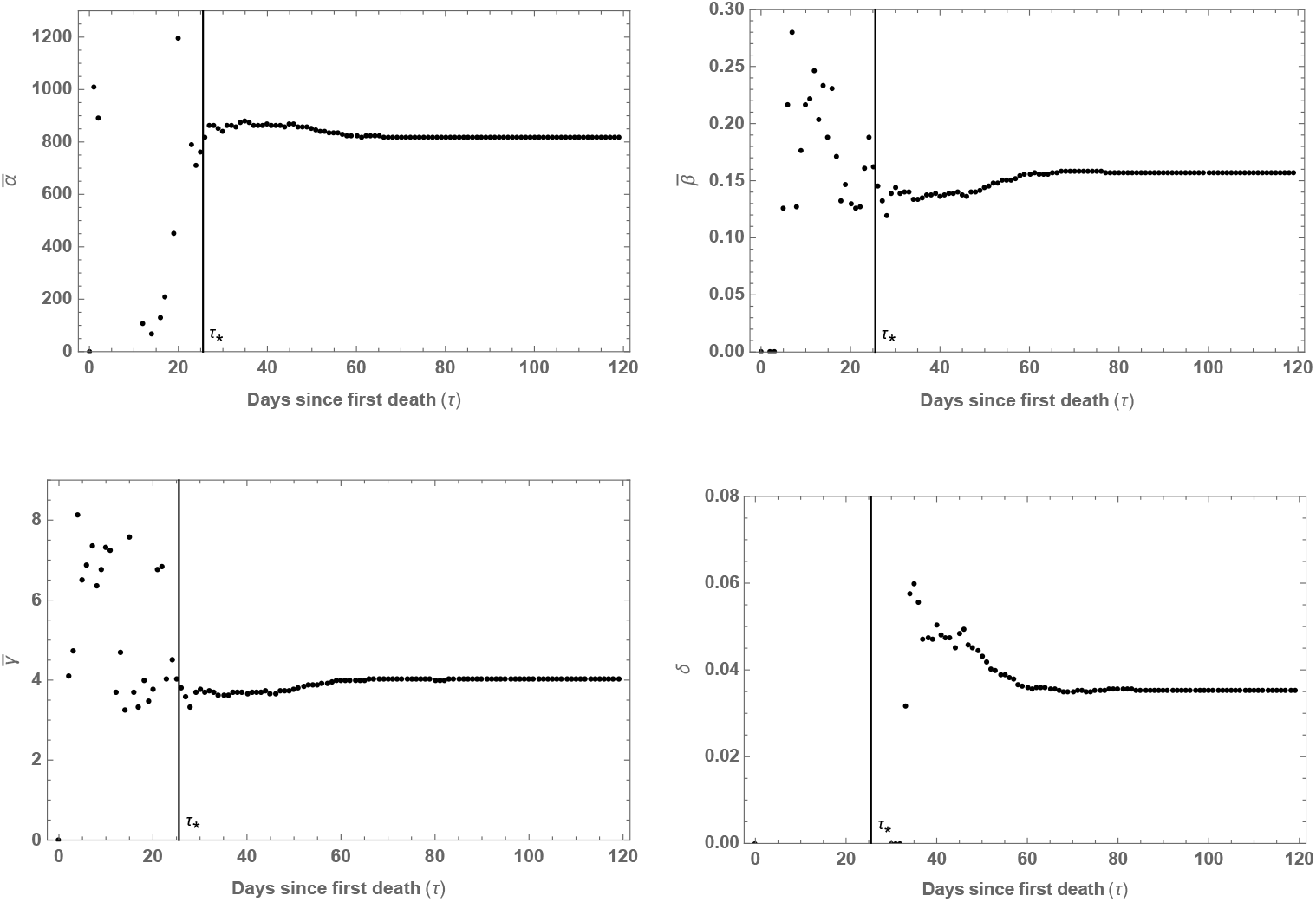
Time dependence of the 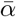, 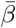, 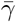 and S constants given by the 4-parameter fit.

## 3 Time evolution of the infection rate discontinuity

Since the values of the evolution parameters stemming from a daily best fit change, the discontinuity in the infection rate 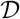 also varies with *τ*. This time evolution will be investigated for different countries starting with the Spanish case, shown in Fig. 10.

**Figure 10:**
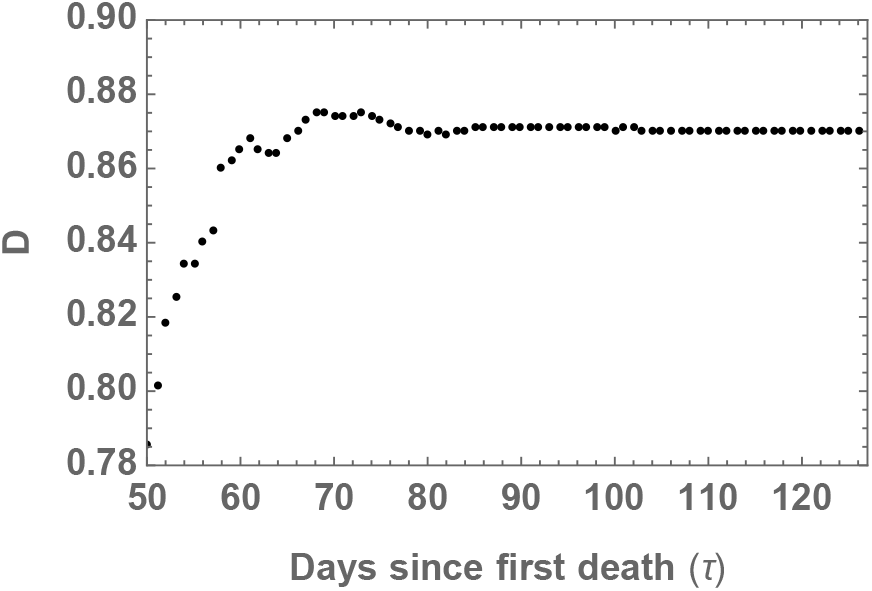
Time dependence of the discontinuity in the infection rate at *τ*_*_ for Spain.

It can be seen that in order to obtain a stable value for 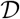 it is needed to enter the stability region of all parameters in the fit, *δ* in particular. To compare among different countries it is needed to define common criteria to fix *τ*_*_ since this variable also carries a time dependence as it is shown in Fig. 11. The chosen procedure will be to calculate the relevant functions at the time (*τ_R_*) when the value of estimated remaining deaths in the pandemia is 2.5 per cent of the total, in this case *τ_R_* = 84.305 (see Fig. 12) and, correspondingly, *τ*_*_ = 25.542.

**Figure 11:**
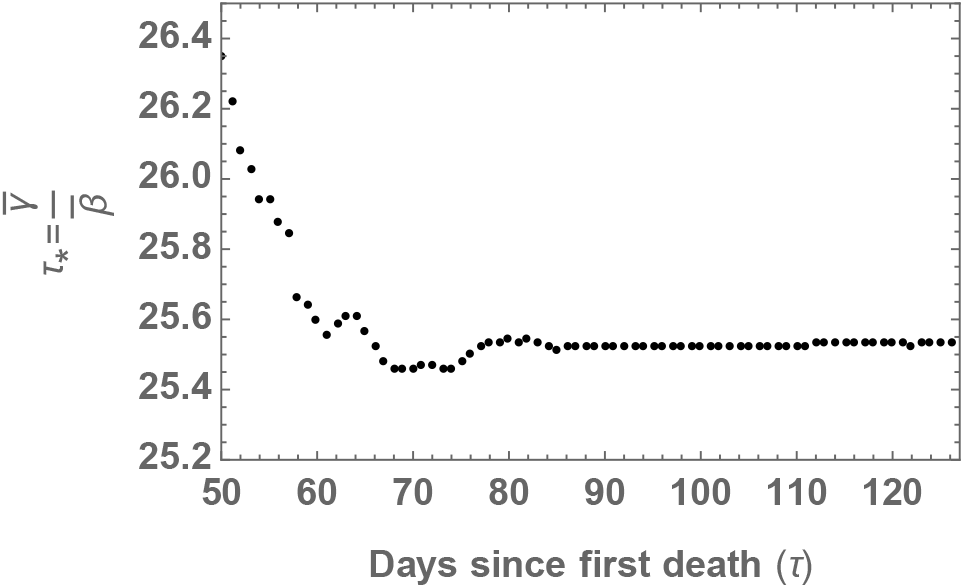
Time dependence of 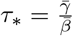 for Spain.

**Figure 12:**
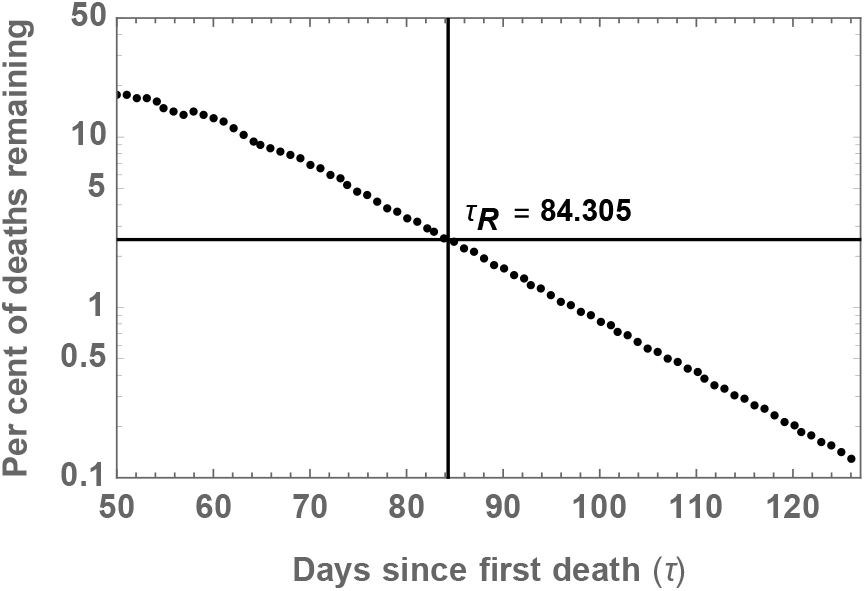
Estimated fraction of remaining deaths for Spain.

This approach will be followed when comparing the data in Spain with those in Germany, Italy and Switzerland. These countries are at a very late stage of their pandemic evolution and the quality of the reported data is high (although it does not account for all the possible deaths due to the pandemic, see, *e.g*., [10], at least there is a clear criterium in the reporting: to have been confirmed by polymerise chain reaction (PCR) or equivalent tests).

The best fit to the data in the four cases has been calculated in Figs. 4, 5, 13, 14 and 15. The associated values for 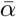, 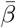, 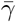 and *δ* are given in Table 1. 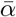 corresponds to the highest value of deaths per day in the curve for *z*′ generated by the fit. The ratio between 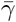 and 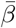, *τ*_*_, corresponds to the position of that maximum and it is shown in Table 2. In this table the total number of casualties (calculated with Eq. (23)) is also presented together with the percentage of deaths taking place after reaching the peak in *z*′, *i.e*., when *τ* > *τ*_*_.

**Table 1:** Best fit values for the four pandemic evolution parameters together with the discontinuity in the time dependence of the infection rate 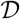 for Spain, Italy, Germany and Switzerland.

| | $\bar{\alpha}$ | $\bar{\beta}$ | $\bar{\gamma}$ | $\delta$ | $\mathcal{D}$ |
| --- | --- | --- | --- | --- | --- |
| Spain | 819.32 | 0.1572 | 4.0131 | 0.0354 | 0.8707 |
| Italy | 747.06 | 0.1057 | 3.5585 | 0.0269 | 0.8480 |
| Germany | 227.74 | 0.0937 | 2.8910 | 0.0343 | 0.7128 |
| Switzerland | 59.491 | 0.1098 | 3.0257 | 0.0420 | 0.6881 |

**Figure 13:**
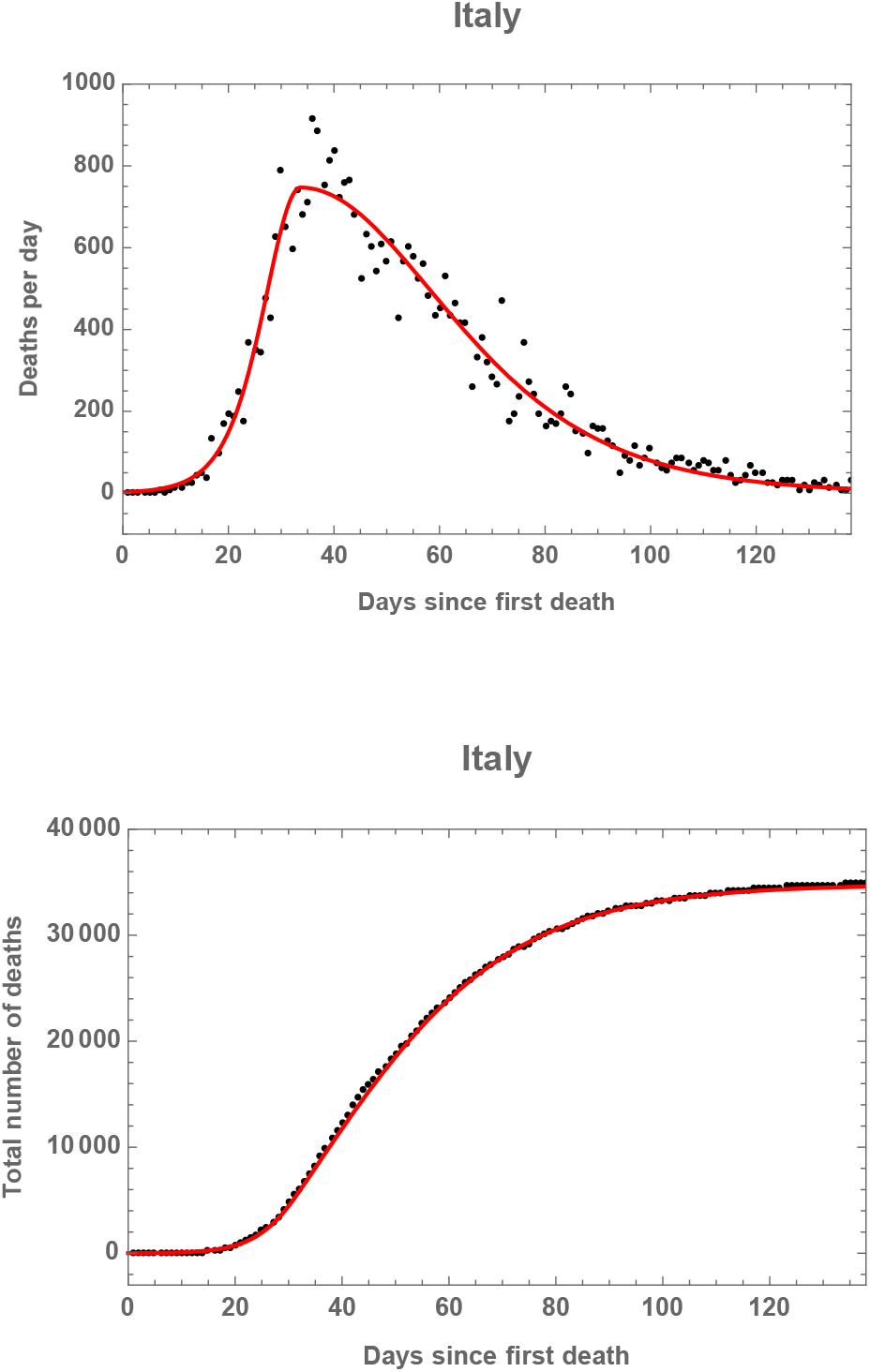
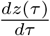and *z*(*τ*) for Italy.

**Figure 14:**
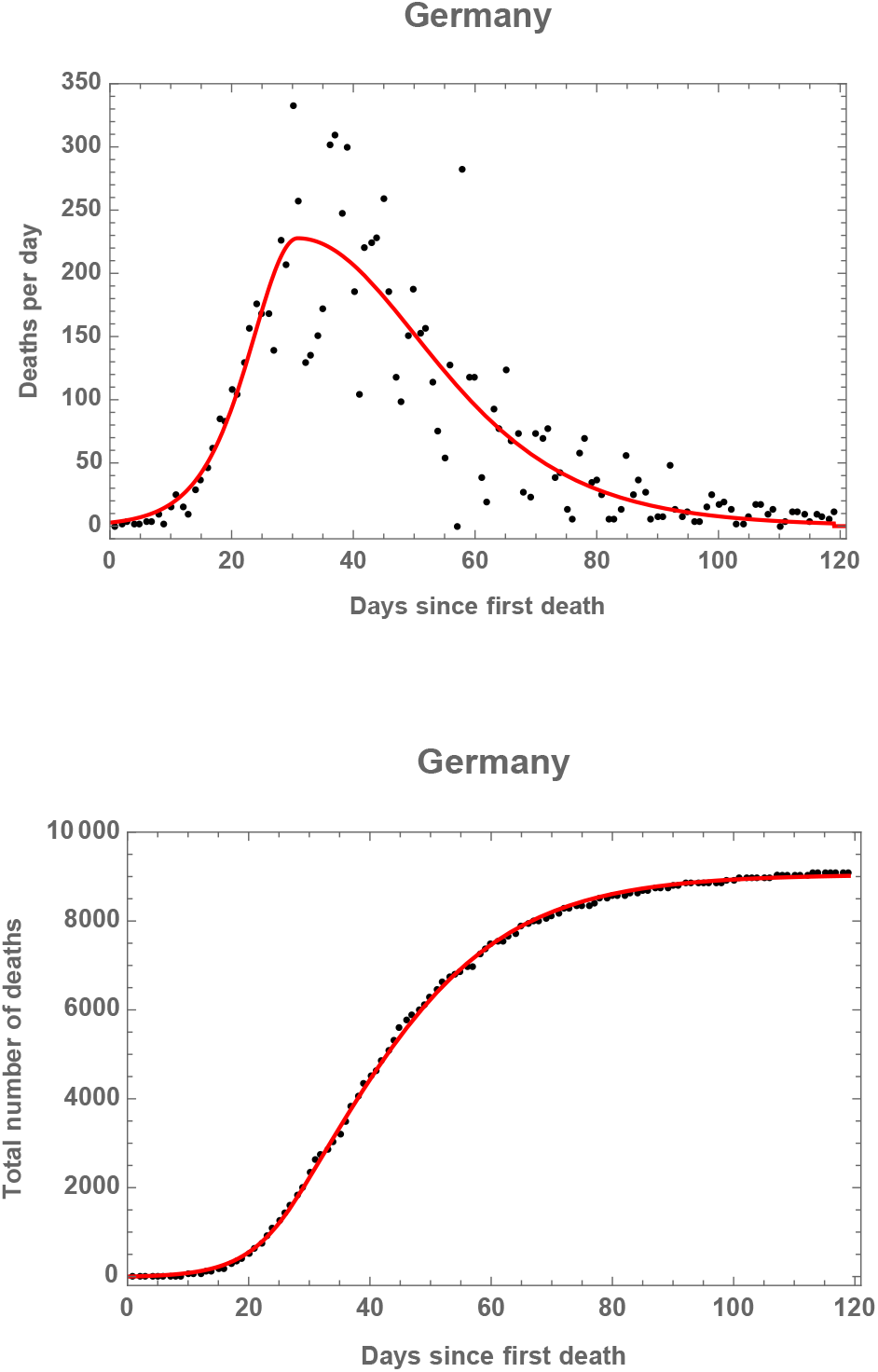
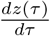 and z(*τ*) for Germany.

**Figure 15:**
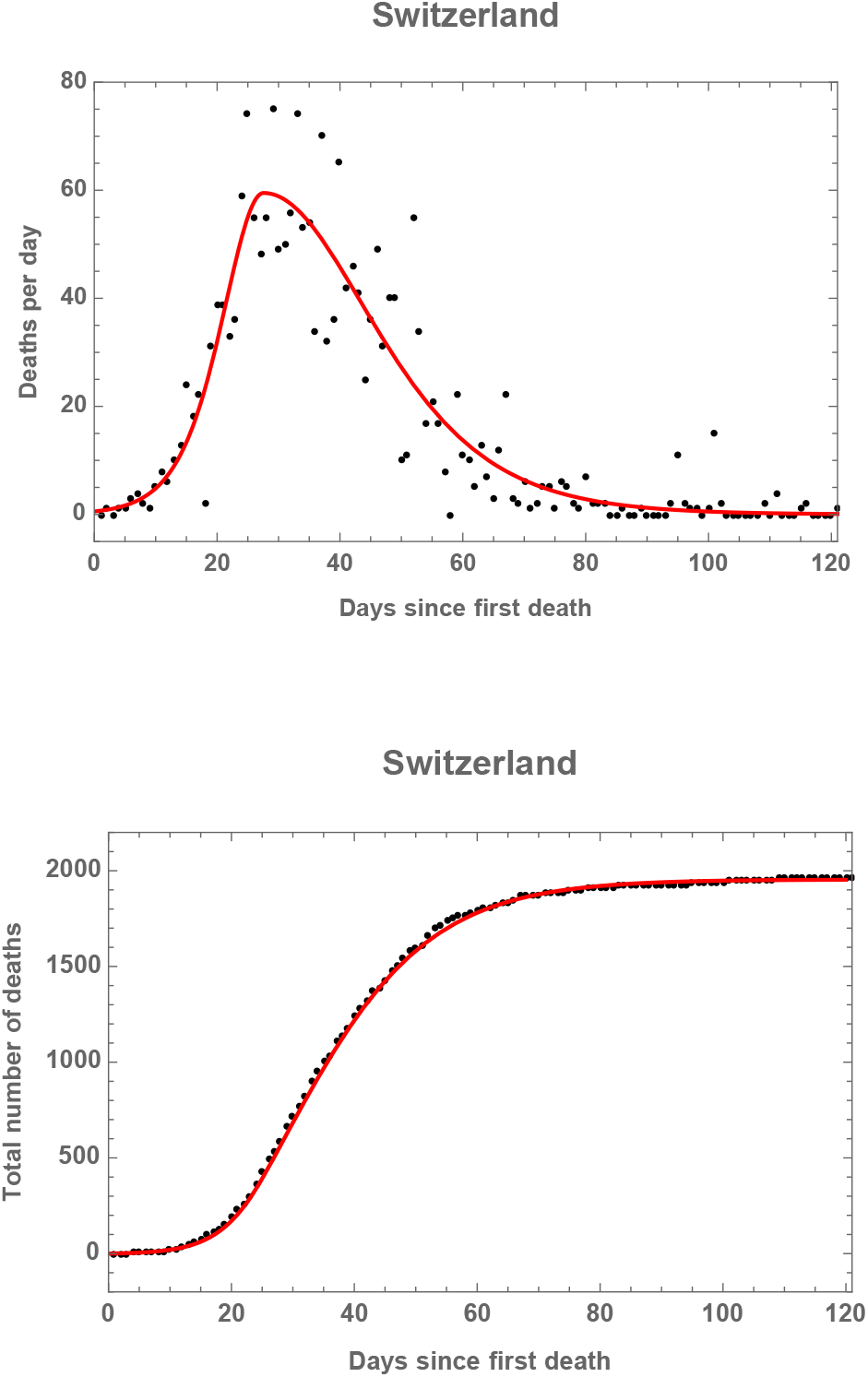
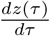 and z(*τ*) for Switzerland.

**Table 2:** Total number of reported casualties (up to July 8^th^ 2020) for Spain, Italy, Germany and Switzerland together with the percentage of deaths after reaching the peak in the number of daily deaths, which takes place at time τ_*_.

| | Total | % after peak | $\tau_*$ |
| --- | --- | --- | --- |
| Spain | 28328 | 81.613 | 25.529 |
| Italy | 34783 | 79.707 | 33.677 |
| Germany | 9046 | 73.290 | 30.864 |
| Switzerland | 1954 | 72.400 | 27.561 |

The main result in this work is to propose 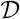, the discontinuity in the time dependence of the infection rate, as a distinct parameter to determine the effectiveness of a particular lockdown in a given country. From its definition in Eq. (38) it can be seen that in an ideal scenario, where the infection rate becomes zero (*ρ*_*_ = 0) after *τ*_*_, 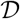 would be equal to one. The values for 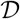 in Spain and Italy, shown in Table 1, are very high, above 0.8. This means that the confinement measures in those countries have been successful. This coincides with the fact that in both the lockdown has been strict. In Germany and Switzerland, where the lockdown has been implemented in a less severe manner, the value for 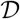 is around 0.7. As explained above, this study has been performed for each country at the date when 97.5 per cent of the total deaths has been reached. This is a convergent region for the discontinuity as shown in Fig. 16 where *τ*_*_ sets the origin of the time coordinate for each nation independently. It is worth noting the correlation between the discontinuity here discussed and the percentage of total casualties taking place after *τ*_*_, which is shown in Table 2. From these results it can be argued that a successful lockdown implies having a larger fraction of casualties after reaching the peak in the *z*′ distribution of deaths per day.

**Figure 16:**
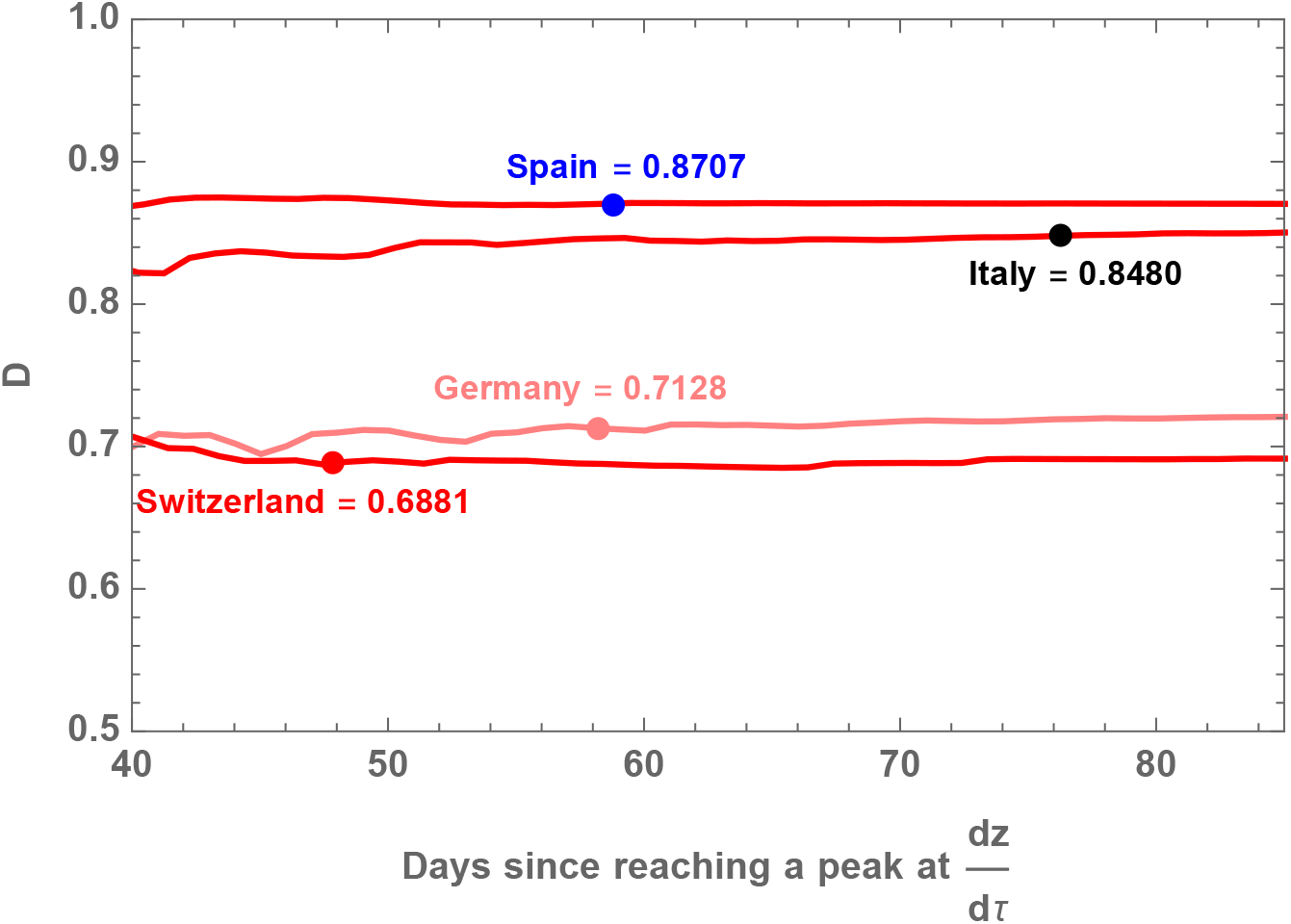
Time dependence for the 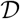 variable. The dots show when 97.5 per cent of the total deaths has been reached in each country. The origin of time is *τ*_*_ for each nation.

To conclude, it is interesting to perform the previous analysis using the available data for the excess in average total mortality instead of the officially reported cases associated to COVID-19. This will be done only for the case of Spain using the data provided by EuroMOMO, the European mortality monitoring activity (https://www.euromomo.eu). The total mortality for all causes is shown in Fig. 17 together with the expected one stemming from an average from previous years.

**Figure 17:**
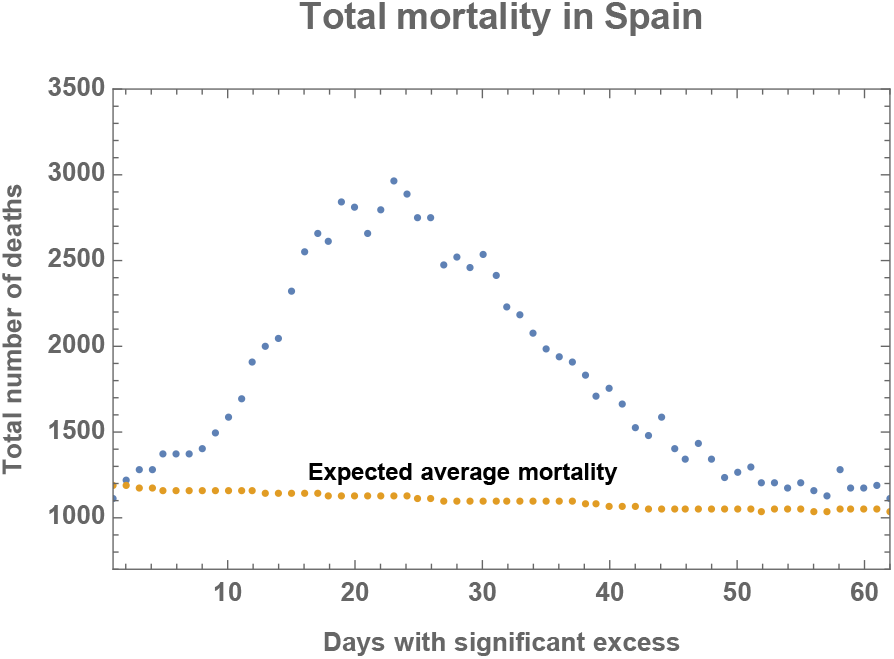
Total mortality in Spain during the first outburst of pandemic evolution in 2020.

Applying Eq. (35), the obtained values for the best fit of the excess in deaths during the first outburst of pandemic evolution are

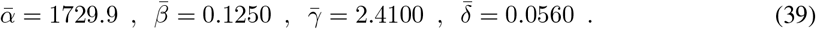

The corresponding curves are shown in Fig. 18. In this case the per cent of deaths after the peak is 69.496 and the value of the discontinuity drops to 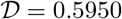 (considering the full data in Fig. 18). The total number of deaths would rise up to 44667. These numbers have to be interpreted with care since there are many factors with influence in the excess of deaths which are not necessarily related to the lockdown measures.

**Figure 18:**
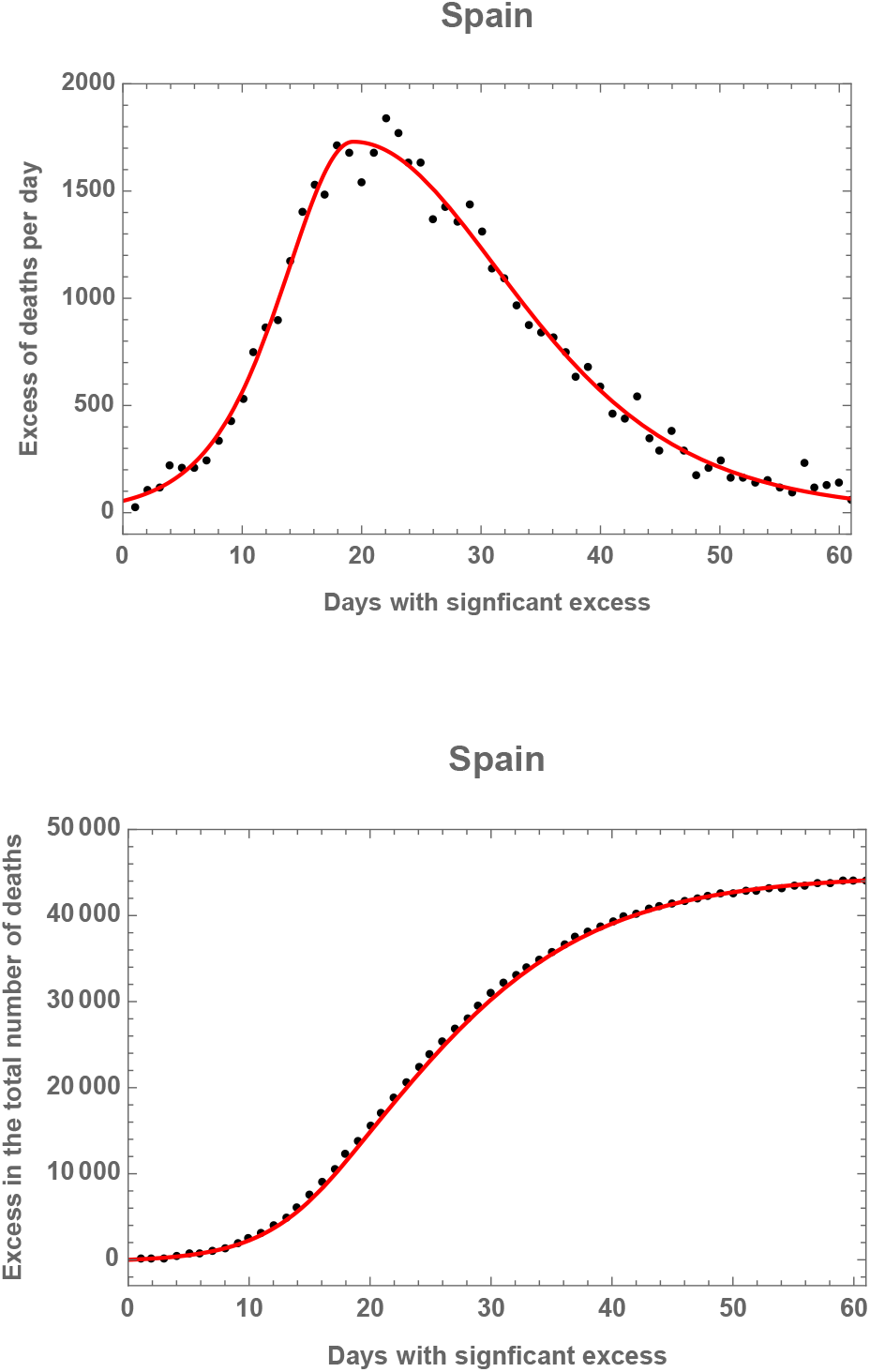
Excess in total mortality in Spain during the first outburst of pandemic evolution in 2020.

## 4 Conclusions

A variation of the original Kermack-McKendrick model for the description of epidemic evolution has been introduced. An asymmetry w.r.t. the maximum in the reported deaths per day is needed in order to describe the transition from a fast rise of the number of deaths at an initial stage of the pandemic to a slow decrease afterwards which is severely affected by the implementation of different lockdown measures within the susceptible population.

This leads to a four parameter model of the evolution of the system which generates an accurate description of it. This point has been proven using the available data for reported fatal infections due to COVID-19 in the case of Spain, Italy, Germany and Switzerland.

The asymmetry in this description generates a discontinuity in the time dependence of the infection rate which has been evaluated in Eq. (38). The closer this variable gets to one, the more efficient the lockdown or confinement measures in a particular country have been. The values for the nations under study have been calculated and are shown in Table 1. **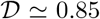** for Spain and Italy, indicating that the lockdown has been more successfully (in terms of reducing the deadly infection rate) applied than in Germany and Switzerland where **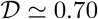**.

This is a general method of study of epidemic evolution which can be easily implemented (a 4-parameter fit to the function in Eq. (35) for the daily number of fatal infections and subsequent evaluation of Eq. (38)) and hopefully can aid in the decision making process associated to the current pandemic situation in any susceptible to infection set of the population.

## Data Availability

The public data considered here have been taken from "Daily New Deaths in Spain" in the real time world statistics generated by Worldometer at https://www.worldometers.info/coronavirus/country/spain/. The same source for officially reported deaths has been used in all the studies of this work.

The public data provided by EuroMOMO, the European mortality monitoring activity https://www.euromomo.eu) have also been used.

## Acknowledgements

I would like to thank Alberto Casas, Grigorios Chachamis, David Gordo, Kiko Llaneras and Douglas Ross for interesting discussions. This work has been supported by the Spanish Research Agency (Agencia Estatal de Investigatión) through the grant IFT Centro de Excelencia Severo Ochoa SEV-2016-0597, and the Spanish Government grants FPA2015-65480-P, FPA2016-78022-P.

## Footnotes

1 The data considered here have been taken from “Daily New Deaths in Spain” in the real time world statistics generated by Worldometer at https://www.worldometers.info/coronavirus/country/spain/. The same source for officially reported deaths has been used in all the studies of this work.

